# Investigating Musculoskeletal Pain Among Pregnant Women in Dhaka Division: A Cross-Sectional Study

**DOI:** 10.1101/2024.04.27.24306482

**Authors:** Jalal Uddin, Shahida Sultana Shumi

## Abstract

**Objectives:** This study aimed to investigate musculoskeletal pain among pregnant women in Dhaka Division, Bangladesh, considering demographic factors, pain and pregnancy-related experiences.

**Materials and Methods:** Data were collected from 300 pregnant women attending outpatient clinical check-ups at two hospitals in Dhaka Division, Bangladesh. A standardized questionnaire covering demographic information, pregnancy-related inquiries, and pain assessment using the Numeric Pain Rating Scale (NPRS) was administered. Data were analyzed using SPSS version 25 software.

**Results:** The majority of participants were aged 20-25 years, resided in rural areas, and had completed primary education. Most participants were housewives. A significant proportion reported experiencing musculoskeletal pain, predominantly in the second trimester. Associations were found between musculoskeletal pain and both age (p=0.025) and trimester of pregnancy (p=0.008).

**Conclusions:** The findings emphasized significant associations between maternal age and musculoskeletal pain, highlighting the importance of considering age-related factors in maternal care. Additionally, a high prevalence of musculoskeletal pain during the second trimester was observed.

## Introduction

The musculoskeletal system is a highly integrated network of bones, muscle groups, tendons, ligaments, and connective tissues that deliver shape to the human body and permit it to move. 1 It supports other structures and gives the structure and stability to walk, carry, grasp, paintings, workout, and perform sports of day-by-day dwelling [1]. Being pregnant causes hormonal and different physiologic changes that cause weight benefit, joint laxity, and sundry biomechanics, all stressors that may result in musculoskeletal pain and initiate harm [2].

Pregnancy-induced biomechanical, hormonal, and vascular changes are likely to contribute to a wide range of musculoskeletal issues [3]. The expanding uterus shifts the body’s center of gravity and exerts mechanical pressure on it [4]. Hormonal fluctuations lead to increased joint laxity in pregnant women. Additionally, fluid retention results in the compression of soft tissues during pregnancy, increasing the risk of musculoskeletal injuries [5]. It has been observed that almost all pregnant women experience some degree of musculoskeletal problems, with some experiencing temporarily disabling symptoms [5].

The primary issue in pregnancy is typically low back pain, with reported occurrences ranging from 30% to 70%. 6 Following this, carpal tunnel syndrome is identified as the second most prevalent musculoskeletal problem [5]. Additionally, muscle cramps, predominantly occurring during sleep and lasting only briefly, constitute another common concern during pregnancy [7].

Spinal ache has been identified as the most prevalent disorder during pregnancy, accompanied by other common issues such as lower and upper extremity pain, muscle cramps, and peripheral neuropathies [8,9]. These musculoskeletal problems have been highlighted as significant contributors to disability and absenteeism among pregnant women [9]. In fact, musculoskeletal issues are estimated to account for about 90% of chronic pain cases, with back pain being particularly prominent [10]. Throughout pregnancy, the female body undergoes significant anatomical and hormonal changes, which may predispose women to various musculoskeletal conditions, increase the risk of injury, or exacerbate pre-existing conditions [10].

The aim of this study was to investigate musculoskeletal pain among pregnant women in Dhaka Division, Bangladesh. Understanding the prevalence and nature of musculoskeletal pain during pregnancy is essential for effective prenatal care. Therefore, our research sought to identify the most common musculoskeletal problems experienced by pregnant women. By examining these issues during routine prenatal checks, healthcare providers can better tailor interventions and support strategies to address the specific needs of pregnant women at different stages of pregnancy.

## Methods

The study collected data from pregnant women attending outpatient clinical check-ups at the Obstetrics and Gynaecology Departments of Dhaka Medical College and Hospital and Zella Hospital Narsingdi in Bangladesh, spanning from April 2019 to September 2019. Employing a convenience sampling technique, a total of 300 pregnant women were included after obtaining written informed consent. The research received approval from the Institutional Review Board (IRB). A formal application was then lodged with the authorities at Dhaka Medical College and Hospital, as well as Zella Hospital Narshingdi, seeking permission to engage clients and utilize their facilities for the study. Following approval, data collection commenced. Data collection occurred via face-to-face interviews, wherein participants responded to a standardized questionnaire covering demographic information, pregnancy-related inquiries, and pain assessment using the Numeric Pain Rating Scale (NPRS). Subsequently, the collected data were analyzed using the SPSS version 25 software program. This investigation was conducted as a cross-sectional study to explore the relationship between demographic factors, pregnancy-related issues, and pain experiences among pregnant women. We performed χ^2^ and t-tests and calculated 95% confidence intervals (CIs) for analyzing the relationship between Patient Characteristics and Musculoskeletal Pain.

## Results

### Patient Characteristics

Among the 300 respondents (see Table-1), the majority of women were aged between 20-25 years (31.3%), followed by those below 20 years (31%), with 27.7% aged between 25-30 years, and the remaining 10% aged over 30 years, with a mean age of 24.27. Regarding living area, data showed that 38.0% were from urban areas, 60.3% from rural areas, and 1.7% from semi-urban areas. Educational qualifications varied, with 1.0% illiterate, 4.7% only able to sign, and 35.3% having completed primary education, followed by 26.7% with secondary education, 15.3% with higher secondary education, 13.7% with a college degree, and 3.3% with other qualifications. Family type indicated that 55.7% lived in extended families, while 44.3% lived in nuclear families. In terms of monthly income, 51.7% earned below 10000 Bangladeshi Taka (BDT), 33.7% earned between 10000-20000 BDT, 10.3% earned between 20000-30000 BDT, and 4.3% earned above 30000 BDT. Regarding occupation, the majority (88.3%) were housewives, 4.3% were students, 1.3% were engaged in government service, 0.7% were teachers, and 5.3% were in other occupations such as nursing or day labor. Religion indicated that 98.7% were Muslim, while 1.3% were Hindu. BMI analysis revealed that 5.0% had a BMI below 18.5, 46.0% had a BMI between 18.5-25, 35.3% had a BMI between 25-30, 13.0% had a BMI between 30-40, and 0.7% had a BMI above 40.

**Table-1.** Patient Characteristics.

| Category | Subcategory | Count (n) | Percentage (%) |
| --- | --- | --- | --- |
| Age | Below 20 | 93 | 31 |
|  | 20-25 | 94 | 31.3 |
|  | 25-30 | 83 | 27.7 |
|  | Above 30 | 30 | 10 |
| Living Area | Urban | 114 | 38 |
|  | Rural | 181 | 60.3 |
|  | Semi-Urban | 5 | 1.7 |
| Educational Qualification | Illiterate | 3 | 1 |
|  | Can sign only | 14 | 4.7 |
|  | Primary Education | 106 | 35.3 |
|  | Secondary Education | 80 | 26.7 |
|  | Higher Secondary Education | 46 | 15.3 |
|  | College Degree | 41 | 13.7 |
|  | Other Education | 10 | 3.3 |
| Family Type | Extended Family | 167 | 55.7 |
|  | Nuclear Family | 133 | 44.3 |
| Family Income | Below 10000 BDT | 155 | 51.7 |
|  | 10000-20000 BDT | 101 | 33.7 |
|  | 20000-30000 BDT | 31 | 10.3 |
|  | Above 30000 BDT | 13 | 4.3 |
| Occupation | Housewife | 265 | 88.3 |
|  | Student | 13 | 4.3 |
|  | Government Service | 4 | 1.3 |
|  | Teacher | 2 | 0.7 |
|  | Other Occupation | 16 | 5.3 |
| Religion | Muslim | 296 | 98.7 |
|  | Hindu | 4 | 1.3 |
| BMI | Below 18.5 | 15 | 5 |
|  | 18.5-25 | 138 | 46 |
|  | 25-30 | 106 | 35.3 |
|  | 30-40 | 39 | 13 |
|  | Above 40 | 2 | 0.7 |

### Pain related Characteristics

The pain-related variables among participants are displayed (see Table-2). A significant majority, constituting 72.0% of participants, reported experiencing pain, while the remaining 28.0% did not. Pain duration varied, with 64.0% experiencing pain for less than 3 months, and 8.0% for more than 3 months. Descriptions of pain nature included 27.7% reporting it as sharp, 35.7% as dull, 1.7% as burning, 5.7% as shooting, and 1.3% as other. Severity, as measured by the NPRS scale, indicated that 24.7% reported mild pain (score <4), 39.3% reported moderate pain (score 4-7), and 8.0% reported severe pain (score >7). Reference of pain was indicated by 23.7%, while 48.3% reported no reference. Specific types of pain, such as headache (10.0%), neck pain (1.7%), arm pain (2.7%), forearm pain (2.7%), hand pain (2.3%), low back pain (38.3%), lower abdominal pain (36.3%), leg pain (18.3%), ankle pain (5.3%), and foot pain (4.3%) were reported. The pattern of pain was described as constant by 10.0% of participants and intermittent by 62.0%.

**Table-2.** Pain related Characteristics.

| **Category** | **Subcategory** | **Count (n)** | **Percentage (%)** |
| --- | --- | --- | --- |
| Pain | Yes | 216 | 72 |
|  | No | 84 | 28 |
| Pain duration | <3 months | 192 | 64 |
|  | >3 months | 24 | 8 |
| Nature of pain | Sharp | 83 | 27.7 |
|  | Dull | 107 | 35.7 |
|  | Burning | 5 | 1.7 |
|  | Shooting | 17 | 5.7 |
|  | Others | 4 | 1.3 |
| Severity of pain | Mild <4 | 74 | 24.7 |
|  | Moderate (4-7) | 118 | 39.3 |
|  | Severe >7 | 24 | 8 |
| Reference of pain | Yes | 71 | 23.7 |
|  | No | 145 | 48.3 |
| Headache | Yes | 30 | 10 |
|  | No | 186 | 62 |
| Neck pain | Yes | 5 | 1.7 |
|  | No | 211 | 70.3 |
| Arm pain | Yes | 8 | 2.7 |
|  | No | 208 | 69.3 |
| Forearm pain | Yes | 8 | 2.7 |
|  | No | 208 | 69.3 |
| Hand pain | Yes | 7 | 2.3 |
|  | No | 209 | 69.7 |
| Low back pain | Yes | 115 | 38.3 |
|  | No | 101 | 33.7 |
| Lower abdominal pain | Yes | 109 | 36.3 |
|  | No | 107 | 35.7 |
| Leg pain | Yes | 55 | 18.3 |
|  | No | 161 | 53.7 |
| Ankle pain | Yes | 16 | 5.3 |
|  | No | 200 | 66.7 |
| Foot pain | Yes | 13 | 4.3 |
|  | No | 203 | 67.7 |
| Pain pattern | Constant | 30 | 10 |
|  | Intermittent | 186 | 62 |

### Pregnancy related Characteristics

This study revealed various pregnancy-related characteristics (see Table-3). 28.0% of respondents were in the 1st trimester, while 45.0% were in the 2nd trimester, and 27.0% were in the 3rd trimester. Regarding complaints during urination, 17.7% of respondents reported pain, while 82.3% did not experience pain. In terms of pregnancy history, 35.3% had a single pregnancy, 23.3% had multiple pregnancies, and 41.3% had no previous pregnancy. Additionally, 26.0% reported a history of miscarriage, and 6.7% reported a history of abortion. Furthermore, 31.7% had a history of normal vaginal delivery, 1.0% had assisted delivery, and 25.7% had a history of cesarean section.

**Table-3.** Pregnancy related Characteristics.

| Category | Subcategory | Count (n) | Percentage (%) |
| --- | --- | --- | --- |
| Trimester | 1st Trimester | 84 | 28 |
|  | 2nd Trimester | 135 | 45 |
|  | 3rd Trimester | 81 | 27 |
| Pain during urination | Yes | 53 | 17.7 |
|  | No | 247 | 82.3 |
| Previous history of pregnancy | Single pregnancy | 106 | 35.3 |
|  | Multiple pregnancies | 70 | 23.3 |
|  | No previous history | 124 | 41.3 |
| Previous history of miscarriage | Yes | 78 | 26 |
|  | No | 222 | 74 |
| History of abortion | Yes | 20 | 6.7 |
|  | No | 280 | 93.3 |
| Previous history of childbirth | Normal vaginal delivery | 95 | 31.7 |
|  | Assisted delivery | 3 | 1 |
|  | Caesarean section | 77 | 25.7 |

### Bivariate Analysis of Patient Characteristics and Musculoskeletal Pain

The bivariate analysis, (see Table-4), yielded several significant findings. Participants reporting musculoskeletal pain exhibited a slightly higher mean age of 24.55 years (SD = 4.64), compared to those without musculoskeletal pain, whose mean age was 23.71 years (SD = 4.63) (p = 0.025). Additionally, no statistically significant difference in participant distribution between urban and rural areas was observed based on musculoskeletal pain status (p = 0.41). Musculoskeletal pain status did not show a significant association with education level (all p > 0.05) or income level (p = 0.858). Similarly, there was no significant difference in musculoskeletal pain status based on occupation (p = 0.863). Although a trend suggesting a difference in family types was noted, it was not statistically significant (p = 0.089). Regarding the trimester of pregnancy, participants in the second trimester had a higher proportion of musculoskeletal pain (48.8%) compared to the first (22.4%) and third trimesters (28.9%) (p = 0.008). Furthermore, no significant associations were found between musculoskeletal pain status and previous history of pregnancy (p = 0.272) or previous history of miscarriage (p = 0.836).

**Table-4:** Bivariate Analysis of Patient Characteristics and Musculoskeletal Pain.

| **Variables** | **Musculoskeletal Pain**<br>**n (%)**<br>**N=201 (67.0)** | **No Musculoskeletal Pain**<br>**n (%)**<br>**N= 99 (33.0)** | **p-value** |
| --- | --- | --- | --- |
| Age, Mean (SD) | 24.55 (4.64) | 23.71 (4.63) | 0.025 |
| **Area** |  |  | 0.410 |
| Urban | 83 (41.3) | 36 (36.4) |  |
| Rural | 118 (58.7) | 63 (63.6) |  |
| **Education** |  |  | 0.453 |
| Illiterate | 9 (4.5) | 8 (8.1) |  |
| Primary | 75 (37.3) | 31 (31.3) |  |
| Under College Degree | 85 (42.3) | 41 (41.4) |  |
| College Degree | 32 (15.9) | 19 (19.2) |  |
| **Income** |  |  | 0.858 |
| <25000 BDT | 182 (90.5) | 89 (89.9) |  |
| >25000 BDT | 19 (9.5) | 10 (10.1) |  |
| **Occupation** |  |  | 0.863 |
| Housewife | 178 (88.6) | 87 (87.9) |  |
| Others | 23 (11.4) | 12 (12.1) |  |
| **Family Types** |  |  | 0.089 |
| Nuclear | 96 (47.8) | 37 (37.4) |  |
| Extended | 105 (52.2) | 62 (62.6) |  |
| **Trimester of pregnancy** |  |  | 0.008 |
| First trimester | 45 (22.4) | 39 (39.4) |  |
| Second trimester | 98 (48.8) | 37 (37.4) |  |
| Third trimester | 58 (28.9) | 23 (23.2) |  |
| **Previous history of pregnancy** |  |  | 0.272 |
| Single | 73 (36.3) | 33 (33.3) |  |
| Multiple | 51 (25.4) | 19 (19.2) |  |
| None of them | 77 (38.3) | 47 (47.5) |  |

| Previous history of miscarriage |  |  | 0.836 |
| --- | --- | --- | --- |
| Yes | 53 (26.4) | 25 (25.3) |  |
| No | 148 (73.6) | 74 (74.7) |  |

## Discussion

In our study, we observed a significant association between age and musculoskeletal pain among pregnant women, indicating that as the age increased, there was a heightened likelihood of experiencing muscular pain. This finding is consistent with a study, who found that increasing maternal age was associated with joint pains and lower back pain (LBP) [11]. Another study reported no significant difference in the age of respondents experiencing musculoskeletal problems during pregnancy [12]. This disparity suggests that while age may influence musculoskeletal pain in some studies, it may not be a significant factor in others.

Our study encompassed 300 pregnant women, with the majority falling within the 20-25 years age range, followed by a significant proportion below 20 years old and between 25-30 years old. This distribution differs slightly from the findings of a study, where the mean age of participants was 30.9 ± 5.0 years, with a significant portion having attained a university-level education. 9 Similarly, a study reported a diverse age range among their sample of pregnant women, with the highest prevalence of musculoskeletal disorders observed in those aged 35-49 years [13].

Our investigation uncovered a diverse educational background among participants, with minimal illiteracy (1%) and significant proportions completing primary (35.3%) and secondary (26.7%) education. Moreover, a notable percentage held a college degree (13.7%), indicating varied socioeconomic profiles within the study population. Regarding income, the study revealed varied levels among participants: 51.7% earned below 10000 BDT monthly, 33.7% fell within the 10000-20000 BDT range, 10.3% earned 20000-30000 BDT, and 4.3% earned over 30000 BDT monthly. In contrast, a study found participants with educational attainment up to the 12th class level, coupled with a relatively higher average family income exceeding 18,000 BDT per month [13]. This income disparity underscores the significance of socioeconomic factors in comprehending the musculoskeletal health of pregnant women, as educational and income differences may impact access to healthcare resources and influence health outcomes.

This study evaluated the mean height, weight, body mass index (BMI) of the participants. Among all respondents (n=15), 5.0% had a BMI below 18.5, while 46.0% fell within the BMI range of 18.5-25. Additionally, 35.3% of respondents had a BMI between 25-30, with 13.0% falling in the range of 30-40, and a minor proportion (0.7%) having a BMI exceeding 40.

A study found that the mean BMI of pregnant women was 24.84 ± 4.62 kg/m2, with 3.2% categorized as low weight, 52.6% as normal weight, 27.5% as overweight, and 16.7% as obese [14]. These findings provide insights into the diverse socioeconomic backgrounds of the study participants, which may influence various health outcomes, including musculoskeletal health during pregnancy.

We found that a higher proportion of participants residing in rural areas reported experiencing musculoskeletal pain compared to those living in urban areas. Specifically, 58.7% of participants from rural areas reported musculoskeletal pain, while only 41.3% of urban residents reported similar pain experiences. These findings are consistent with previous research, which also observed a higher prevalence of musculoskeletal pain among urban dwellers compared to rural communities. For instance, urban residents comprised approximately 59% of the study group, while rural residents constituted around 41%. 15 Moreover, musculoskeletal pain was more prevalent among urban participants (54.4%) compared to their rural counterparts (45.6%) [13].

This study revealed varying degrees of musculoskeletal pain among participants, with 64.0% experiencing acute pain (<3 months) and 8.0% reporting chronic pain (>3 months). Specific discomforts included headache (10.0%), low back pain (38.3%), and lower abdominal pain (36.3%). A study found that 50.7% of pregnant participants experienced symptoms, with low back pain (55.6%), ankle pain (25.9%), and knee pain (16.6%) being the most prevalent [13].Common musculoskeletal complaints during pregnancy, including low back pain (70.7%) and hip pain (32.1%), particularly in the third trimester [9]. These findings underscore the diverse nature of musculoskeletal discomfort during pregnancy and emphasize the need for targeted interventions to alleviate maternal discomfort and improve overall well-being.

Common musculoskeletal problems across trimesters, with lower back pain, pelvic girdle pain, and carpal tunnel syndrome prevalent in the first trimester; lower back pain, cramps, and pelvic girdle pain in the second trimester; and lower back pain, cramps, and pedal edema in the third trimester [6]. In our study, participants in the second trimester exhibited a higher prevalence of musculoskeletal pain (48.8%) compared to the first (22.4%) and third trimesters (28.9%) (p = 0.008). Notably, calf pain, low back pain, and pelvic girdle pain were the most commonly reported dysfunctions during the second trimester [16]. A meta-analysis reported a global prevalence of back pain in the first, second, and third trimesters as 28.3%, 36.8%, and 47.8%, respectively [17]. These findings underscore the dynamic nature of musculoskeletal discomfort throughout pregnancy, emphasizing the importance of tailored interventions for pregnant individuals across different stages of gestation.

In our study, a similar pattern of childbirth methods was observed, with 31.7% of participants having a history of normal vaginal delivery, 25.7% undergoing cesarean section, and 1.0% experiencing assisted delivery. This aligns with a finding, where among 550 mothers, 74.2% had vaginal deliveries, while 25.8% underwent cesarean deliveries [18].

Our study provides insights into the multifaceted influences on musculoskeletal health during pregnancy, encompassing factors such as age, education, income, and trimester-specific discomforts. Recognizing these complexities underscores the necessity for comprehensive maternal care strategies, targeted interventions, and further investigation to address socioeconomic disparities and trimester-specific pain. However, several limitations should be noted. Firstly, the study was funded personally, potentially limiting the extent of data collection and analysis. Secondly, time constraints restricted the ability to obtain a larger sample size, potentially impacting representativeness. Lastly, the study included only 300 participants, possibly limiting generalizability.

## Conclusions

Our study observed a significant association between maternal age and musculoskeletal pain, emphasizing the need to consider age-related factors in maternal care strategies. Furthermore, our findings highlight the diverse socioeconomic backgrounds of pregnant women, underscoring the importance of tailored interventions that address socioeconomic disparities. We also identified trimester-specific discomforts, with a notable prevalence of musculoskeletal pain during the second trimester. Understanding these trimester-specific challenges is crucial for developing targeted interventions to alleviate maternal discomfort.

## Data Availability

All data produced in the present study are available upon reasonable request to the authors

## Author contributions

To write: Jalal Uddin

Literature research: Shahida Sultana Shumi

Statistical analysis: Jalal Uddin

Data collecting: Shahida Sultana Shumi

